# Health Impacts of Micro- and Nanoplastics in Humans: Systematic Review of *In Vivo* Evidence

**DOI:** 10.1101/2025.07.10.25331209

**Authors:** Hien Anh Tran Anna, Daniel Hengyi Tang, Cai Ting Yong, John Joson Ng, Andrew Fu Wah Ho

## Abstract

**Background:** Microplastics and nanoplastics (MNPs; <5 mm) are pervasive environmental contaminants with potential human health implications. Although laboratory models implicate MNPs in oxidative stress, inflammation, and endocrine disruption, a comprehensive synthesis of direct *in vivo* human evidence is lacking. We aimed to systematically review studies measuring MNPs in living human subjects and summarize associated health findings.

**Methods:** We systematically searched PubMed, Web of Science, Scopus, Cochrane and Embase through 26 December 2024. Two investigators independently screened and selected original research articles that quantified MNPs in biological samples from living humans (blood, tissues, or fluids). We excluded animal, *in vitro*, cell-line, and injection-based studies, as well as reports on non-plastic micro- and nanoparticles. Data extraction, performed in duplicate, included study design, participant characteristics, detection methods, polymer types, and reported health outcomes. Methodological quality was appraised using Risk Of Bias in Non-Randomized Studies - of Exposures (ROBINS-E). The primary outcome was the presence and burden of MNPs; secondary outcomes were clinical or biomarker associations. We did not perform meta-analysis due to heterogeneity.

**Findings:** From 5 522 records, 25 studies (n=1498 participants) met inclusion. Studies employed pyrolysis-gas chromatography/mass spectrometry (n=9), Raman (n=8), infrared spectroscopy (n=7), and Fourier-transform infrared spectroscopy (n=3), often combined with microscopy. Predominant polymers were polyethylene, polypropylene, polyvinyl chloride, polyethylene terephthalate, and polystyrene. In cardiovascular research (five studies; n=454), higher thrombus and plasma MNP burdens correlated with inflammatory markers and adverse cardiac events. Reproductive research (seven studies; n=327) linked semen and tissue MNP levels to reduced sperm quality and higher burdens in tumor and placental samples. Gastrointestinal research (nine studies; n=537) associated fecal MNPs with liver enzyme elevations and gut dysbiosis. Respiratory (three studies; n=171) and ocular (one study; n=49) research detected MNPs in airway fluids and vitreous humor, with suggestive links to inflammation and increased intraocular pressure. ROBINS-E assessments indicated moderate to high risk of confounding and exposure-measurement bias; consistency across detection modalities was limited.

**Interpretation:** Human *in vivo* evidence confirms that MNPs accumulate in multiple organ systems and are associated with inflammation and functional impairment. Methodological heterogeneity and bias constrain causal inference. Standardized, prospective cohort studies with rigorous exposure assessment and confounder control are needed to advance understanding and guide policy.

**Funding:** This study was funded by the SingHealth Duke-NUS Academic Medical Centre under the Nurturing Clinician Scientist Award scheme (15/FY2025/P1/17-A45) and the Clinician Investigator Advancement Programme (15/FY2022/CIVA/03-A03).

## INTRODUCTION

Microplastics and nanoplastics (MNPs; <5 mm) have emerged as urgent environmental and public health challenges (1). They arise from the fragmentation of larger plastic waste (single-use packaging, synthetic textiles) and intentionally manufactured microparticles in personal care and pharmaceutical products (2). During the COVID-19 pandemic, widespread use of disposable masks, gloves, and personal protective equipment exacerbated plastic pollution, amplifying environmental MNP loads (3,4). Owing to their small size and chemical persistence, MNPs readily infiltrate ecosystems, enter food chains, and accumulate in the human body (5,6). Their ubiquity in modern life, coupled with their ability to interact with biological systems at the cellular level, has prompted increasing interest in their potential impact on human health.

Historically, research on MNPs has relied on animal and *in vitro* studies to postulate their potential effects on human health (7,8). These studies suggest that MNPs may induce oxidative stress, inflammation, cytotoxicity, and disrupt endocrine and immune systems (9–13). Studies done *in vivo* have largely focused on measuring exposure levels to MNPs in blood, stool and other tissues without establishing causal relationships between MNP exposure and clinical health outcomes (14–17). This limits our ability to provide targeted, evidence-based public health guidance. From a policy-making perspective, there is a risk of either underestimating potential harms or inciting undue fear among the public.

Recently, more human-centric, *in vivo* studies analysing the relationship between MNPs on human health have emerged, yet no systematic effort has synthesized these nascent findings. To address this gap, we performed a systematic review of all original research investigating the impact of MNPs in human subjects. Our objectives were to (1) catalogue the types, sources, and detection methods of MNPs identified *in vivo*, (2) summarize associated health outcomes across organ systems, and (3) critically appraise methodological strengths and limitations. By clarifying the current state of human-centric evidence, we aim to inform future research priorities and provide a foundation for evidence-based policy.

## METHODS

### Search Strategy and Selection Criteria

We performed a systematic review of original research articles available on PubMed, Web Of Science, Scopus and Embase databases from inception to 26 December 2024. The search strategy was developed under the guidance of a medical information specialist. The review was carried out according to the Preferred Reporting Items for Systematic Reviews and Meta-Analysis (PRISMA) 2020 guidelines, and two researchers independently reviewed and approved the articles for their full-text eligibility (18). The complete search strategy can be found in Supplementary Table 1.

We considered all original research articles that investigated the effect of MNPs on human health. Studies were included if they analysed tissue, blood or organ samples from live human subjects only. No restrictions were imposed on the health outcomes, year of publication or language. We did not consider research that was performed on animals, organoids, lab-engineered tissue models, or human cell lines. Studies artificially introducing MNPs into the human body were also excluded, as were those that focused on non-plastic materials bound to the surface of MNPs.

Where data was available, we analyzed the most common sources of MNP exposure. We also reviewed the methods used by the articles for detecting, quantifying MNPs in human tissues and evaluating their health impact based on the framework below (Table 1).

**Table 1:** Analytical framework for *in vivo* human microplastic studies. A structured overview of key components—quality control, identification, quantification, localisation, characterisation, and biological impact—each paired with the central question addressed and the recommended methods or tools.

| Component | Key Questions | Methods/Tools |
| --- | --- | --- |
| Quality control | Are the results accurate and reproducible? | Reference materials (e.g., from NIST), standardised protocols, whole blank controls, determination of limit of detection/quantification, interlaboratory validation, contamination prevention |
| Identification | What types of plastics are present? | Raman, FTIR, LDIR, Py-GC/MS |
| Quantification | How much plastic is present? | Flow cytometry, microscopy ($\mu$), Py-GC/MS, LDIR, μ-FTIR |
| Localisation | Where are the plastics located in the tissue? | μ-Raman, μ-FTIR, SEM |
| Characterisation | What are the physical properties of the plastics? | Microscopy |
| Biological impact | How does the presence of plastics impact human health outcomes? | Toxicology assays, biomarker analysis, validated tools appropriate to the organ, system, or disease assessed |

### Data Analysis

The risk-of-bias assessment was performed using ROBINS-E, a tool developed specifically for non-randomized studies of exposure in the context of systematic reviews of observational epidemiological studies (19). The ROBINS-E score constitutes seven domains that evaluate risk of bias of a study secondary to confounding, measurement of exposure, selection of participants, post-exposure interventions, missing data, measurement of outcome and selection of reported results. Each domain is classified into one of four categories: “Low”, “Some Concerns”, “High” and “Very High”. An overall judgement score is also noted.

### Role of the Funding Source

The funders of the study had no role in study design, data collection, data analysis, data interpretation or writing of the report.

## RESULTS

Our search identified 5522 unique records. After removing duplicates, we screened 4 527 titles and abstracts, retrieved 81 full texts, and ultimately included 25 articles (Figure 1).

**Figure 1.**
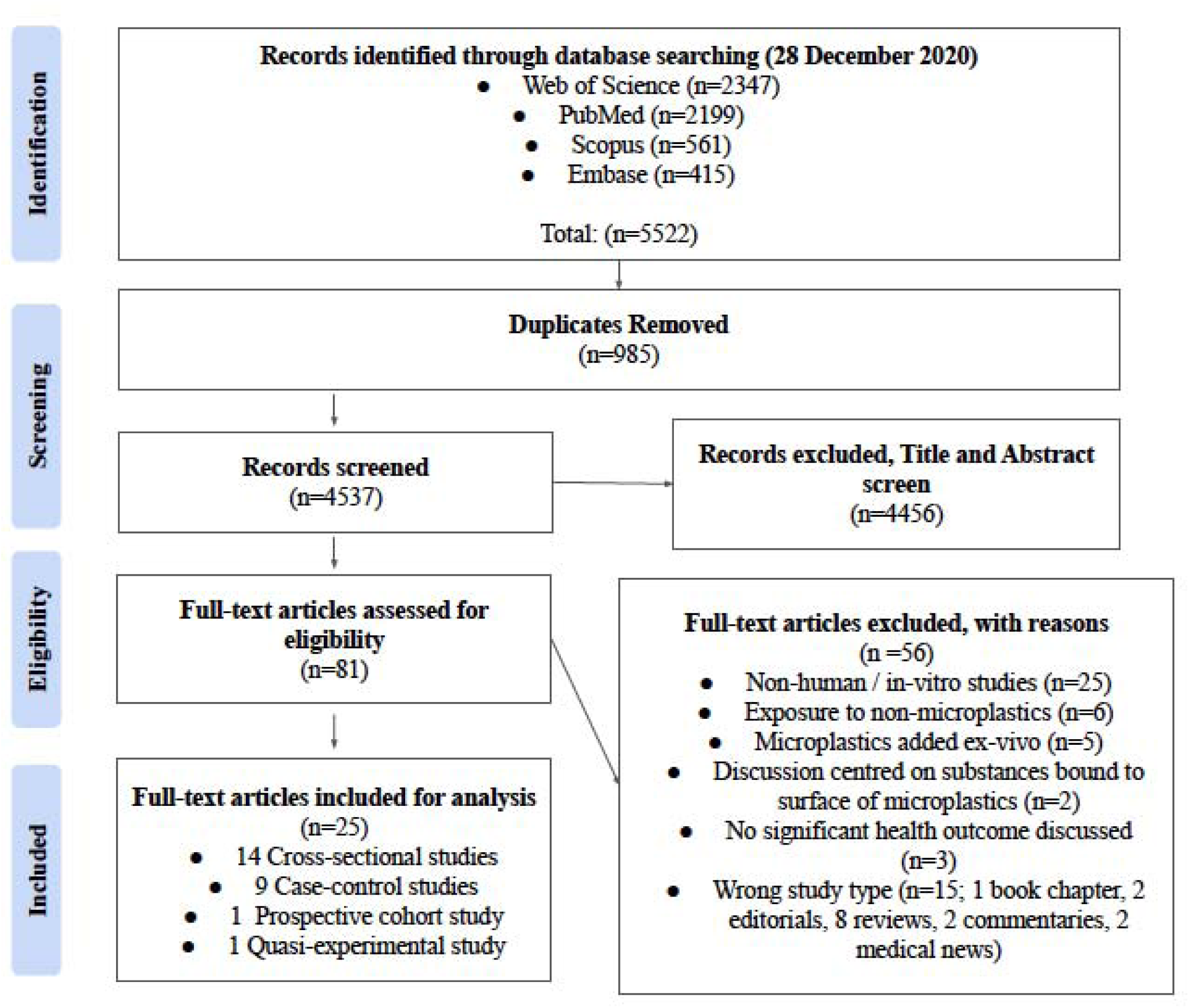
PRISMA 2020 flow diagram of study selection. Flow of records through the systematic review process. Database searches of Web of Science, PubMed, Scopus, and Embase (through 28 December 2020) yielded 5 522 records, of which 985 duplicates were removed. After title and abstract screening of 4 537 records, 81 full-text articles were assessed for eligibility. Fifty-six were excluded (non-human or in vitro studies, non-microplastic exposures, ex vivo additions, surface-bound substance analyses, lack of clinical outcomes, or inappropriate publication types). Twenty-five studies met inclusion criteria for qualitative synthesis (14 cross-sectional, nine case–control, one prospective cohort, and one quasi-experimental).

Of these 25 studies (14–16,20–41), 14 were cross-sectional, nine were case–control, one was quasi-experimental, and one was a prospective cohort. The studies analyzed span a publication window from November 2022 to May 2024. Geographically, the research is predominantly concentrated in China (16 studies), followed by Turkey (three studies), and single contributions from Italy, Iran, Indonesia, Pakistan, Canada, South Korea, and the UK. Seven studies recruited healthy volunteers; the remainder focused on patient groups (e.g., acute coronary syndrome, intra-uterine growth restriction (IUGR) pregnancies, chronic rhinosinusitis) (Table 2).

**Table 2.**
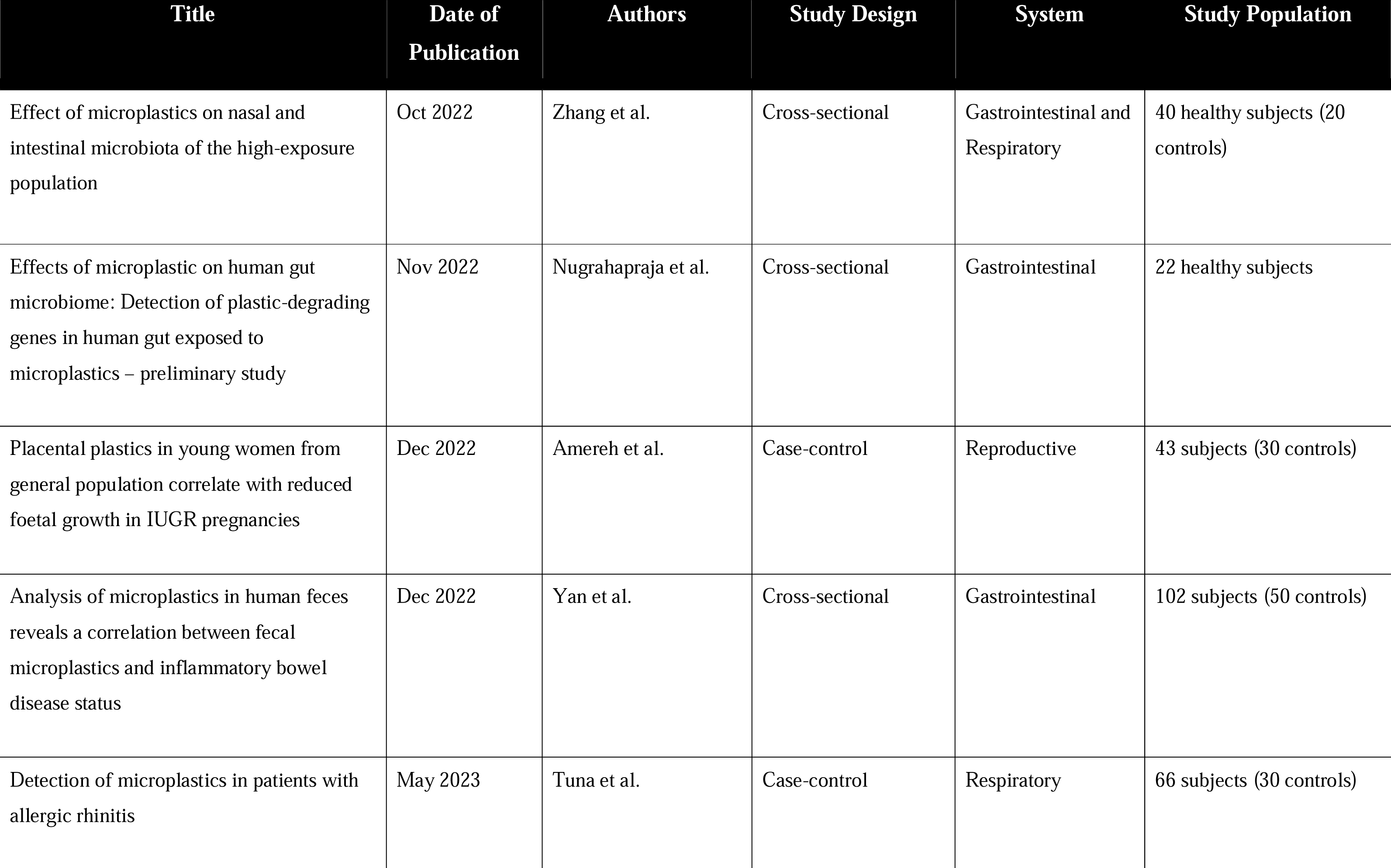
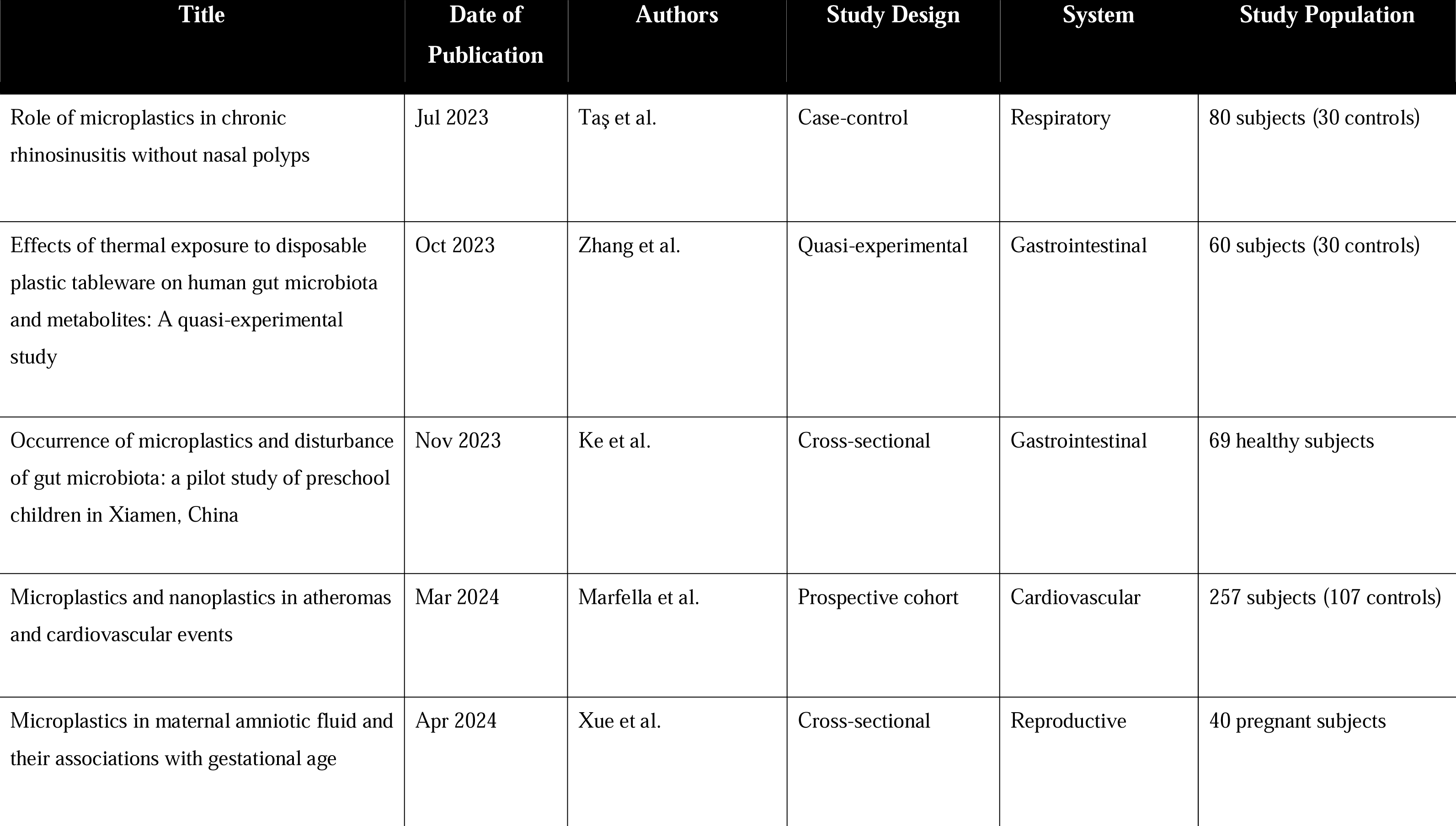
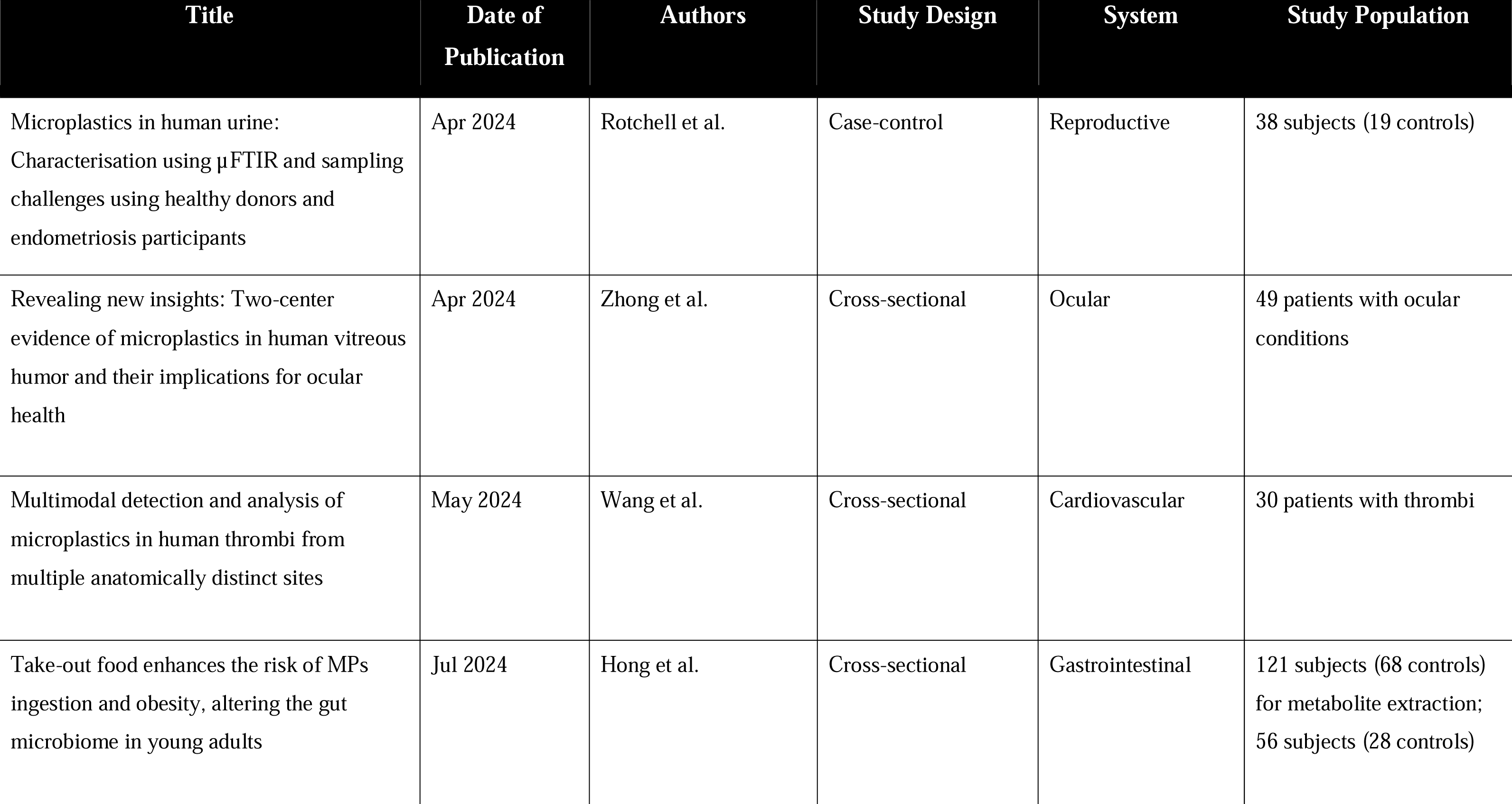
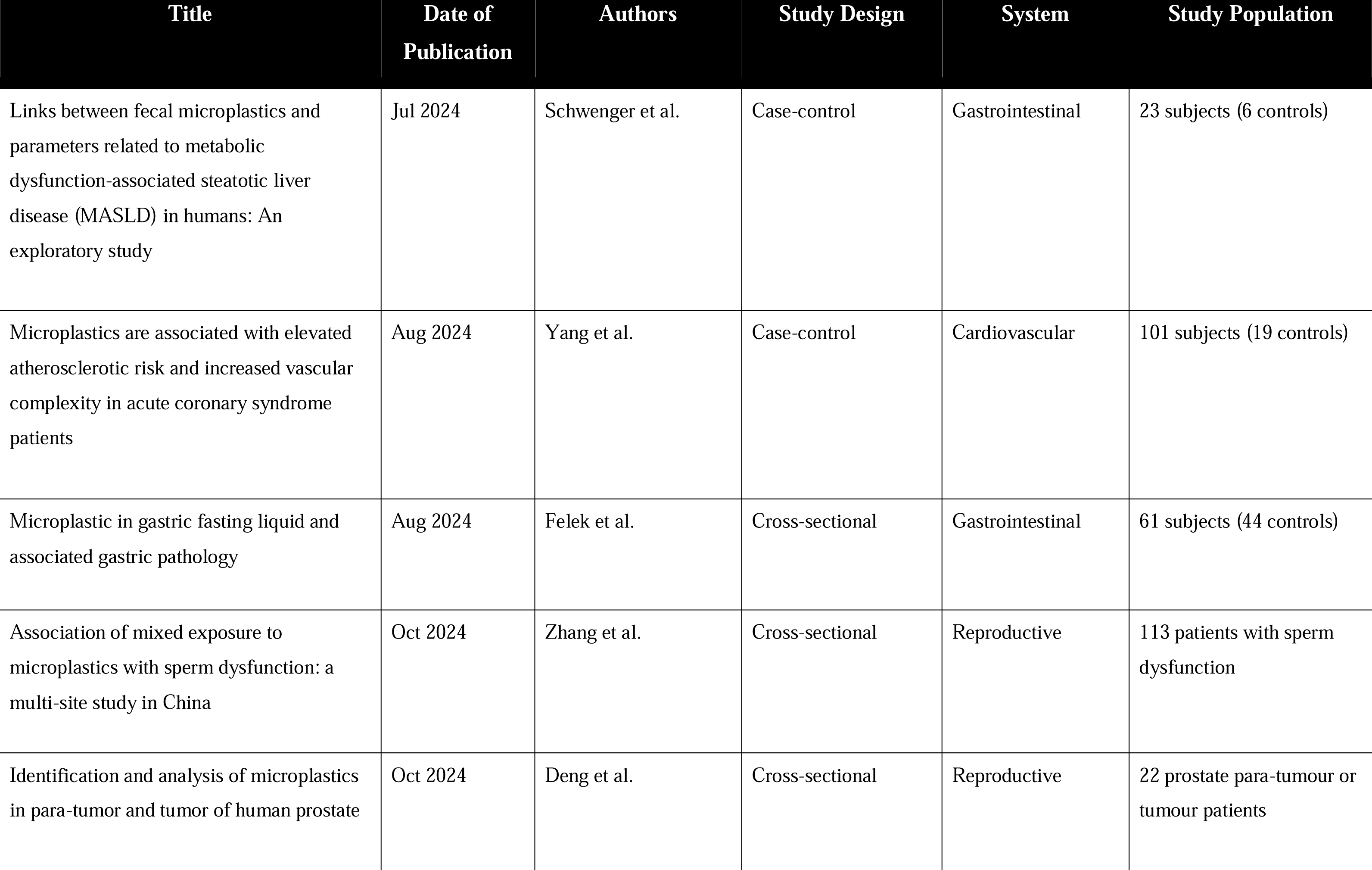
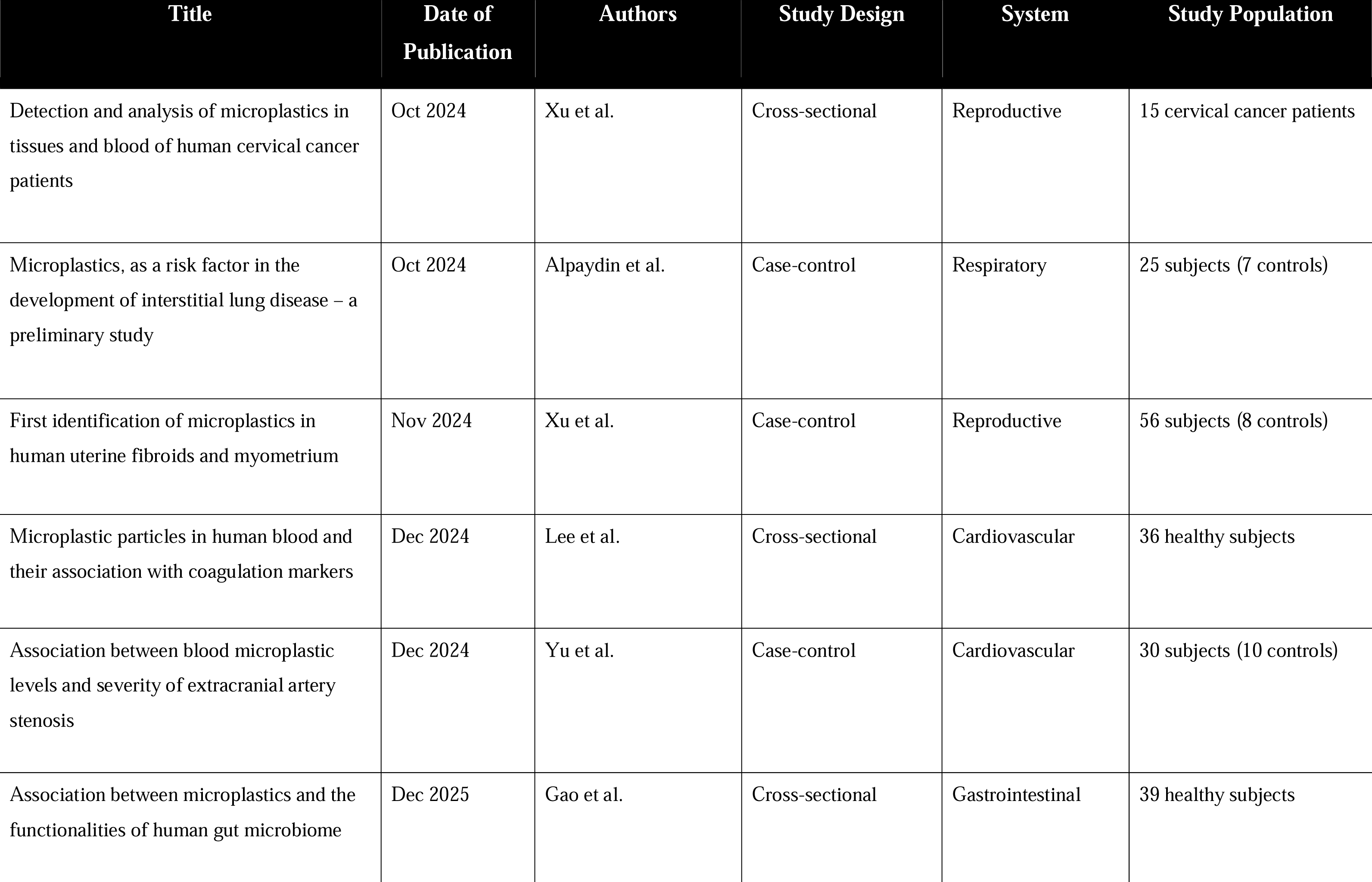
List of the 25 studies included in this systematic review, showing key details: publication date, authors, study design, organ system investigated, and study population characteristics. Abbreviations. : ACS = Acute Coronary Syndrome; DVT = Deep Vein Thrombosis; ECA = Extracranial Artery; IBD = Inflammatory Bowel Disease; ILD = Interstitial Lung Disease; IUGR = Intrauterine Growth Restriction; LD-IR = Laser Direct Infrared; MASLD = Metabolic Dysfunction-Associated Steatotic Liver Disease; MI = Myocardial Infarction; MNPs = Microplastics and Nanoplastics; MPs = Microplastics; Py-GC/MS = Pyrolysis-Gas Chromatography/Mass Spectrometry; SEM = Scanning Electron Microscopy; TEM = Transmission Electron Microscopy; μ-FTIR = micro-Fourier Transform Infrared Spectroscopy.

| Title | Date of Publication | Authors | Study Design | System | Study Population |
| --- | --- | --- | --- | --- | --- |
| Effect of microplastics on nasal and intestinal microbiota of the high-exposure population | Oct 2022 | Zhang et al. | Cross-sectional | Gastrointestinal and Respiratory | 40 healthy subjects (20 controls) |
| Effects of microplastic on human gut microbiome: Detection of plastic-degrading genes in human gut exposed to microplastics – preliminary study | Nov 2022 | Nugrahapraja et al. | Cross-sectional | Gastrointestinal | 22 healthy subjects |
| Placental plastics in young women from general population correlate with reduced foetal growth in IUGR pregnancies | Dec 2022 | Amereh et al. | Case-control | Reproductive | 43 subjects (30 controls) |
| Analysis of microplastics in human feces reveals a correlation between fecal microplastics and inflammatory bowel disease status | Dec 2022 | Yan et al. | Cross-sectional | Gastrointestinal | 102 subjects (50 controls) |
| Detection of microplastics in patients with allergic rhinitis | May 2023 | Tuna et al. | Case-control | Respiratory | 66 subjects (30 controls) |
| Role of microplastics in chronic rhinosinusitis without nasal polyps | Jul 2023 | Taş et al. | Case-control | Respiratory | 80 subjects (30 controls) |
| Effects of thermal exposure to disposable plastic tableware on human gut microbiota and metabolites: A quasi-experimental study | Oct 2023 | Zhang et al. | Quasi-experimental | Gastrointestinal | 60 subjects (30 controls) |
| Occurrence of microplastics and disturbance of gut microbiota: a pilot study of preschool children in Xiamen, China | Nov 2023 | Ke et al. | Cross-sectional | Gastrointestinal | 69 healthy subjects |
| Microplastics and nanoplastics in atheromas and cardiovascular events | Mar 2024 | Marfella et al. | Prospective cohort | Cardiovascular | 257 subjects (107 controls) |
| Microplastics in maternal amniotic fluid and their associations with gestational age | Apr 2024 | Xue et al. | Cross-sectional | Reproductive | 40 pregnant subjects |
| Microplastics in human urine: Characterisation using μFTIR and sampling challenges using healthy donors and endometriosis participants | Apr 2024 | Rotchell et al. | Case-control | Reproductive | 38 subjects (19 controls) |
| Revealing new insights: Two-center evidence of microplastics in human vitreous humor and their implications for ocular health | Apr 2024 | Zhong et al. | Cross-sectional | Ocular | 49 patients with ocular conditions |
| Multimodal detection and analysis of microplastics in human thrombi from multiple anatomically distinct sites | May 2024 | Wang et al. | Cross-sectional | Cardiovascular | 30 patients with thrombi |
| Take-out food enhances the risk of MPs ingestion and obesity, altering the gut microbiome in young adults | Jul 2024 | Hong et al. | Cross-sectional | Gastrointestinal | 121 subjects (68 controls) for metabolite extraction;<br>56 subjects (28 controls) |
| Links between fecal microplastics and parameters related to metabolic dysfunction-associated steatotic liver disease (MASLD) in humans: An exploratory study | Jul 2024 | Schwenger et al. | Case-control | Gastrointestinal | 23 subjects (6 controls) |
| Microplastics are associated with elevated atherosclerotic risk and increased vascular complexity in acute coronary syndrome patients | Aug 2024 | Yang et al. | Case-control | Cardiovascular | 101 subjects (19 controls) |
| Microplastic in gastric fasting liquid and associated gastric pathology | Aug 2024 | Felek et al. | Cross-sectional | Gastrointestinal | 61 subjects (44 controls) |
| Association of mixed exposure to microplastics with sperm dysfunction: a multi-site study in China | Oct 2024 | Zhang et al. | Cross-sectional | Reproductive | 113 patients with sperm dysfunction |
| Identification and analysis of microplastics in para-tumor and tumor of human prostate | Oct 2024 | Deng et al. | Cross-sectional | Reproductive | 22 prostate para-tumour or tumour patients |
| Detection and analysis of microplastics in tissues and blood of human cervical cancer patients | Oct 2024 | Xu et al. | Cross-sectional | Reproductive | 15 cervical cancer patients |
| Microplastics, as a risk factor in the development of interstitial lung disease – a preliminary study | Oct 2024 | Alpaydin et al. | Case-control | Respiratory | 25 subjects (7 controls) |
| First identification of microplastics in human uterine fibroids and myometrium | Nov 2024 | Xu et al. | Case-control | Reproductive | 56 subjects (8 controls) |
| Microplastic particles in human blood and their association with coagulation markers | Dec 2024 | Lee et al. | Cross-sectional | Cardiovascular | 36 healthy subjects |
| Association between blood microplastic levels and severity of extracranial artery stenosis | Dec 2024 | Yu et al. | Case-control | Cardiovascular | 30 subjects (10 controls) |
| Association between microplastics and the functionalities of human gut microbiome | Dec 2025 | Gao et al. | Cross-sectional | Gastrointestinal | 39 healthy subjects |

12 studies mentioned specific routes of exposure to MNPs for their study population. In particular, takeaway food packaging (n=7) and increased bottled water intake (n=6) was the most significant. Other sources included seafood intake, dairy/formula milk, and occupational exposures among plastic-manufacturing workers.

The main techniques used to identify and quantify MNPs were pyrolysis-gas chromatography mass spectrometry (Py-GC/MS) (n=9), Raman spectroscopy (n=8), laser directed infrared (LDIR) spectroscopy (n=7), and Fourier transform infrared (FTIR) spectroscopy (n=3). 18 studies extended their analysis to characterise and/or localise the MNPs found via microscopy, where the sizes (n=18), shapes (n=15), colours (n=8) of the particles, and their locations in the tissue (e.g., tumor vs para-tumor) were analysed. Two studies further analysed potential correlations between MNPs in the blood and the relevant organ tissue.

ROBINS-E assessment showed that out of the 25 studies, 11 studies were rated “Low Risk,” eight had “Some Concerns,” and six were “High Risk” (Figure 2). We found that the highest risk-of-bias arose from bias due to confounding (D1), followed by bias due to measurement of exposure (D2), and bias in selection of reported results (D6). Bias due to post-exposure interventions (D4) contributed the least to overall judgement (Figure 3).

**Figure 2.**
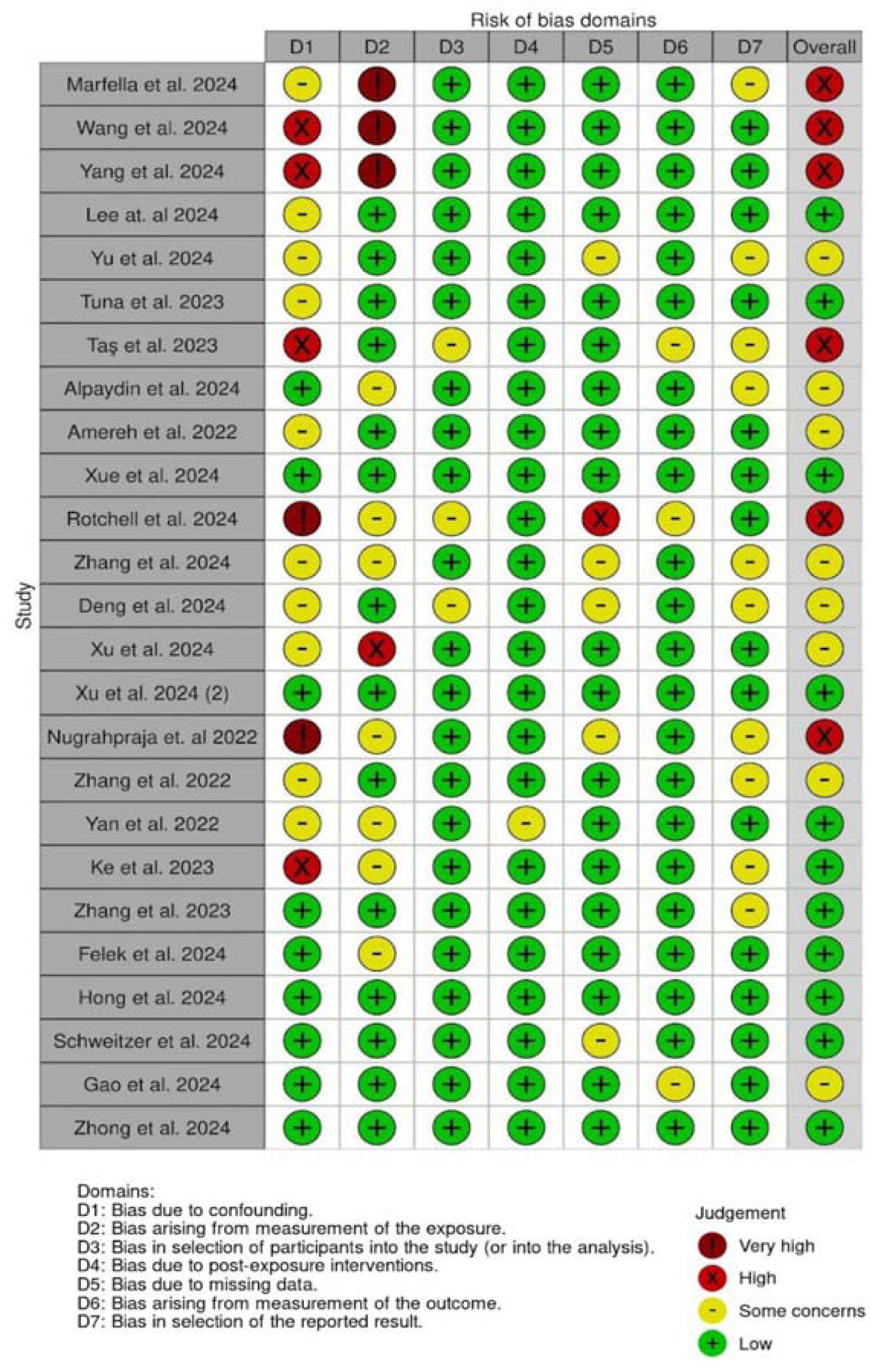
Risk-of-bias assessment of included studies using ROBINS-E. Heatmap summarising risk-of-bias judgments across seven ROBINS-E domains for each of the 25 human *in vivo* studies. Domains are: D1, confounding; D2, exposure measurement; D3, participant selection; D4, post-exposure interventions; D5, missing data; D6, outcome measurement; and D7, selective reporting. Symbols indicate domain-level risk: very high risk (dark red); high risk (red); some concerns (yellow); low risk (green). The “Overall” column shows each study’s maximum domain rating, guiding interpretation of methodological robustness.

**Figure 3.**
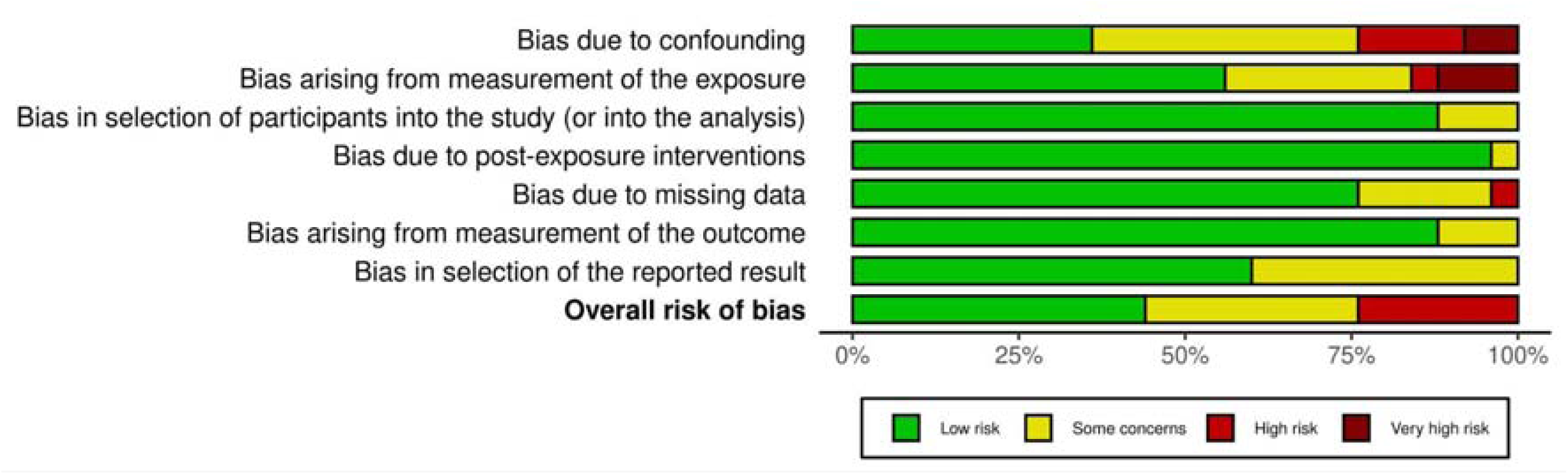
Summary of risk-of-bias across studies by ROBINS-E domain. Stacked bar charts showing the proportion of included human *in vivo* studies rated as low risk (green), some concerns (yellow), high risk (red), and very high risk (dark red) within each ROBINS-E domain: D1 (confounding), D2 (exposure measurement), D3 (participant selection), D4 (post-exposure interventions), D5 (missing data), D6 (outcome measurement), D7 (selective reporting), and the overall risk of bias. Percentages along the x-axis reflect the share of studies falling into each risk category per domain.

## DISCUSSION

MNP pollution has emerged as a global environmental and public health concern, with accumulating evidence suggesting its potential role in adverse health outcomes. Research has predominantly focused on the gastrointestinal, reproductive, and respiratory systems, with more recent investigations exploring potential cardiovascular effects. However, data on the impact of MNPs on other physiological systems remain limited. The field is relatively understudied, and study sizes have generally been small, which may reflect challenges in recruitment, exposure assessment, and methodological standardization. Most studies utilized a cross-sectional design, where the data is collected at a single point in time, and is thus unable to capture changes over time or establish clear causal relationships. Notable exceptions were Marfella et al. and Zhang et al. which employed a longitudinal study design.

The majority of papers show that MNPs can be found in most biological compartments in humans. The most common polymers detected are polyethylene (PE), polypropylene (PP), polyvinyl chloride (PVC), polyethylene terephthalate (PET), and polystyrene (PS), which are widely used in the textiles, consumer goods, and food packaging industries (42–45). Their environmental persistence and propensity to fragment into micro- and nanoscale particles (<10 µm), combined with hydrophobic surfaces and high surface-to-volume ratios, enhance dispersion in air and water, facilitate cellular uptake, and promote systemic bioaccumulation (45). One must also note that FTIR and Raman spectroscopic libraries are most comprehensive for PE, PP, PVC, PET, and PS, making them easier to identify compared to less-studied plastics (46).

Plastic food containers and bottled water appeared to be the most significant source of MNP exposure in our review. One study found MNPs in all samples of polypropylene takeaway containers from seven Chinese cities, ranging from three to 43 particles per container, with PP being the most prevalent polymer (47). Another study of 23 Iranian bottled water brands reported an average of 1,496·7 ± 1,452·2 particles per litre (48). MNPs from plastic packaging are known to leach into food and beverages in the setting of high temperatures, increased ultraviolet exposure, and bacterial activity, and are readily engulfed by macrophages—suppressing lysosomal activity and potentially impairing immune function (49,50). Given the widespread reliance on plastic food packaging and bottled water, these exposure pathways represent a significant public health concern.

Interestingly, although seafood consumption was frequently cited during screening as a primary exposure route (51–53), only one of the 25 included studies identified it as a contributing factor— highlighting an area that requires further investigation. Other exposure pathways mentioned include dairy and formula milk consumption, dust inhalation and occupational exposures. As exposure pathways are complex and often unavoidable, understanding the health effects of MNPs is essential to designing effective public health interventions.

### Impact of MNP exposure on the cardiovascular system

Five of the 25 reviewed articles focus on the effects of MNPs on the human cardiovascular system. Lee et al. (2024) reported PS, PP, PE and PET as the most detected MNPs, while Yang et al. (2024) and Marfella et al. (2024) highlighted PE and PVC as predominant, especially in patients with acute coronary syndrome (ACS). Similarly, Yu et al. (2024) found PVC and polyamide (PA66) were the most abundant MNPs, and Wang et al. (2024) confirmed the presence of PVC, PE and PA66 in thrombi. The ubiquity of PE and PVC may be attributed to their ability to translocate into the bloodstream more easily due to their size and chemical composition, hence settling in vascular lesion sites (54).

The presence of MNPs appears to be associated with increased cardiovascular risk. Marfella et al. (2024) demonstrated a correlation between PE levels and inflammatory markers. Yu et al. (2024) purported that MNPs may induce immune cell-associated inflammatory responses in acute coronary syndrome patients. This is consistent with other studies that elaborated upon mechanisms such as the activation of neutrophils by MNPs (55), inducing an immunometabolic active state in macrophages (56). Additionally, the study by Lee et al. (2024) further found associations between elevated MNP levels and alterations in coagulation markers (e.g, longer activated partial thromboplastin time, higher levels of fibrinogen and high-sensitivity C-reactive protein), which are indicators of inflammation and prothrombotic conditions. In vitro studies support these findings, showing polystyrene nanoplastics disrupt fibrinogen structure and promote clot formation Animal studies also demonstrated enhanced thrombus formation triggered by nanoplastics(57–59). Notably, Wang et al. (2024) found MNPs embedded in thrombi, particularly PVC, PE and PA66. This suggests possible physical entrapment of MNPs during thrombus formation (60). Beyond inflammation and thrombosis, MNPs may also exacerbate cardiovascular disease severity. Marfella et al. (2024) reported that patients with MNPs in carotid artery plaques had an increased risk of myocardial infarction, stroke or death. Similarly, Yang et al. (2024) found that increased MNP levels correlated positively with higher SYNTAX scores, indicating more complex coronary artery disease. Yu et al. (2024) also found a positive association between the external carotid artery stenosis percentage and MP concentration (27,33,40).

### Impact of MNP exposure on the reproductive system

Seven of the 25 studies examined MNPs in the human genitourinary system: two in males and five in females. In males, Zhang et al. (35) detected PS, PVC, PE, and PP in semen and urine, with higher concentrations of PC and PP in semen suggesting testes accumulation (61–64).

Polytetrafluoroethylene (PTFE) exposure correlated with reduced sperm count, concentration, and motility, while PVC specifically impaired motility (64). Although murine studies have investigated mechanisms of testicular toxicity, corroborating human data remain scarce (65–67). Deng et al. (36) identified the same four polymers in prostate tumor and adjacent tissues, finding PS, PE, and PVC more abundant in tumors—a pattern mirrored in other organs (15,68,69). Proposed drivers of tumor□specific accumulation include enhanced angiogenesis, proliferation, and pinocytosis, though causality in humans remains to be tested (70).

In females, three studies addressed gynecological conditions. Rotchell et al. (29) found no difference in urinary MNP levels between endometriosis patients and controls, but did not assess MNPs within ectopic lesions; since MNPs can carry endocrine□disrupting chemicals (EDCs) implicated in endometriosis, further tissue□level analyses are warranted (71,72). Xu et al. (15,37,73) consistently reported PE, PP, and PE-co-PP as dominant in cervical cancer, uterine fibroids, and myometrial samples, aligning with other reports of variable MNP profiles (74–79). However, heterogeneity in polymer types and a limited number of human studies preclude firm conclusions. Two studies focused on obstetric outcomes. Xue et al. (28) measured PE, copolymer (CPE), and polyurethane (PU) in amniotic fluid, establishing a negative correlation with gestational age. Amereh et al. (22) showed PS and PE in normal and IUGR placentas, with additional PET and PP in IUGR cases, echoing earlier findings (80–82). A murine model demonstrated a 12% fetal weight reduction after high□dose PS exposure (83), suggesting potential developmental risks, though real□world exposure levels and precise localisation within placental compartments require clarification.

Given that infertility affects 10–15% of couples globally (84,85) and that hormone-sensitive conditions—cervical cancer, IUGR, and prostate cancer—remain major public health burdens (86,87), the pervasive presence of MNPs in reproductive tissues underscores the urgent need for longitudinal, multigenerational studies with standardized exposure assessment and detailed tissue-specific localisation.

### Impact of MNP exposure on the gastrointestinal system

Nine studies have linked gastrointestinal MNP exposure with pathologies like metabolic-associated steatotic liver disease (MASLD), inflammatory bowel syndrome (IBS), obesity risk, and neuropsychiatric disorders such as depression and anxiety.

Subjects with higher MNP exposure tend to exhibit prolonged gastric pathology compared to those with lower exposure, likely due to a chronic proinflammatory state. Schwenger et al. showed an increase in macrophages, killer T cells and natural killer cells in patients with higher exposure to MNPs (32). The pro-inflammatory potential of MNPs has been attributed to several mechanistic pathways. First, MNPs have been shown to induce oxidative stress and neurotoxic effects, thereby disrupting cellular homeostasis (89). Second, they can compromise intestinal barrier integrity, which may facilitate translocation of luminal antigens and further propagate inflammatory responses (90). Third, MNPs have been observed to modulate cytokine expression by upregulating pro-inflammatory mediators such as interleukin-1 alpha (IL-1α) while down regulating anti-inflammatory cytokines (91). These mechanisms collectively contribute to sustained intestinal inflammation, as supported by findings from Schwenger et al. and Felek et al.

The gut microbiome has been shown according to many recent studies to have an impact on many aspects of gut health such as nutrient extraction, metabolism and immunity (86,87,92,93). Dysbiosis—marked by reduced diversity, altered core taxa, or diminished gene richness—may occur when this balance is disrupted. . MNPs have been proposed to impact gut health by damaging the intestinal barrier, inducing apoptosis, altering mucin expression, and triggering inflammation, based on both *in vivo* and *in vitro* studies (94). The effect of MNPs found *in vivo* shows that the degree of disruption is debatable. For example, Zhang et al. observed a significant reduction in alpha diversity with increased MNP load, whereas Nugrahapraja et al. did not find any significant changes on the overall diversity of the gut microbiome. Certain genera such as *Ruminococcus, Dorea, Fusobacterium,* and *Coprococcus* which have shown a correlation with IBD, were found more abundantly.

Separately, Zhang et al. observed that MNP exposure correlates with altered abundances of gut microbial phyla associated with neuropsychiatric conditions—specifically increased *Actinobacteria* and *Proteobacteria* and decreased *Firmicutes* and *Bacteroidota*. The gut-brain axis may mediate these effects, influencing neurological function and mood regulation pathways.

### Impact of MNP exposure on the respiratory system

We observed positive correlation between increased MNP exposure and the occurrence of chronic rhinosinusitis without nasal polyps and allergic rhinitis (23,24). Both upper airway diseases are linked to atopy, characterised by airway hyperresponsivity, eosinophilia and chronic inflammation (23,95,96). MNP exposure is also associated with nasal microbiome dysregulation, notably a reduction in the immunomodulating *Bacteroides spp.* (98). Of note, no dose-response relationship was seen between increased MNP load and the severity of the subjects’ symptoms, as defined by the Score for Allergic Rhinitis (SFAR) and Nasal Obstruction Symptom Evaluation (NOSE). Thus, it is unclear whether reducing MNPs will decrease bothersome symptoms and increase quality of life for such patients.

Alpaydin et al. explored the increased MNP burden in bronchial alveolar lavage (BAL) in patients with interstitial lung disease (ILD) (38). Patients had negative blood MNP levels, pointing away from hematogenous spread and towards inhalation from the environment as the main source of exposure. MNPs have been shown to induce pulmonary fibrosis by promoting epithelial cell ferroptosis and activating oxidative stress signaling pathways in mice models, but no research has been conducted on humans to date (99,100).

Although numerous studies have examined MNP effects in lung models, mice, and human lung cells, few have investigated live human subjects. Consequently, the mechanisms of MNP-induced lung damage remain unclear. Further research should explore potential links with common respiratory conditions such as pneumonia, asthma, and chronic obstructive pulmonary disease.

### Impact of MNP exposure on ocular health

One paper discussed the impacts of MNPs on ocular health (30). The most frequently detected MNPs in the vitreous humour was PA66, followed by PVC and PS. To date, this is the only study analysing the presence of MNPs in the posterior chamber of the eye. Zhong et al. noted positive correlations between increased vitreous MNP burden and increased intraocular pressure (IOP) and aqueous opacities. Increased IOP is closely linked to glaucoma, irreversible optic neuropathy and progressive blindness (101,102). Aqueous opacities can cause significant light scattering before reaching the retina, and patients may complain of poor vision, floaters and dark spots (103,104). In this study, all patients had pre-existing ocular diseases, and any additional pathology can significantly impact their visual function and quality of life.

While Zhang et. al. study gives us a glimpse into the potential consequences of MNP exposure on ocular health, the data is far from sufficient to make any concrete judgement. Future research should focus on exploring the impacts of intravitreal MNPs on ocular markers in healthy subjects. There is also much room to explore the impacts of MNPs on human ocular surface toxicity and anterior chamber pathologies.

### Quality Assessment

#### D1: Bias due to confounding

In our review, we identified bias due to confounding to be the domain that poses the highest risk of compromising the link between MNP exposure and human health outcomes. Notably, seven out of nine studies focusing on the health impact of MNPs on the gastrointestinal system failed to control for subjects’ dietary habit, body mass and exercise habits, all well-established risk factors for GI pathologies (105). Additionally, many studies did not address the role that genetics and family history play in the development of disease states, most notably neoplasms and autoimmune disorders (106).

#### D2: Bias arising from measurement of exposure

There is considerable heterogeneity in detection techniques across—and even within—organ systems, which undermines reliable cross□study comparisons. The most frequently used methods—Raman spectroscopy, FTIR spectroscopy, LDIR, and Py-GC/MS—vary in spatial resolution and detection limits. Table 3 provides an overview of each method’s principles, strengths, and limitations (80,107–112); detailed comparisons appear in other reviews (113–116). Crucially, these techniques require rigorous, tissue□specific validation to eliminate biological matrix interference: for example, a recent Py-GC/MS study (112) found persistent blood-derived contaminants despite extensive digestion, underscoring this need.

**Table 3.** Comparison of analytical methods for microplastic and nanoplastic detection in human samples. Overview of four common spectroscopic and pyrolytic techniques—Raman spectroscopy, FTIR spectroscopy, LDIR spectroscopy, and Py-GC/MS—detailing each method’s underlying principle, quantification capabilities, key advantages, and primary disadvantages.

| Method | Principle | Quantification | Advantages | Disadvantages |
| --- | --- | --- | --- | --- |
| Raman spectroscopy | Measures inelastic light scattering to produce a molecular “fingerprint” spectrum. | **Particle Count:** Yes – provides information on individual particle size, shape, and count.<br>**Mass Concentration:** Indirect – mass estimates require additional calibration and assumptions. | **High spatial resolution:** Can detect particles as small as ~360 nm (107). | **Fluorescence interference:** Biomolecules and pigments may obscure the signal.<br>- Weak and variable signal intensity makes quantitative measurements challenging (108). |
| FTIR spectroscopy | Detects infrared light absorption by molecular bonds, generating characteristic spectra. | **Particle Count:** Yes – provides particle counts and size distribution (typically for particles >20 µm).<br>**Mass Concentration:** Indirect – mass estimates can be derived from particle dimensions using calibration models. | **Non-destructive:** Preserves particle morphology and count data. | **Detection limit:** Generally limited to detecting particles >20 µm (109).<br>- Labor-intensive sample preparation and need for experienced operators. |
| LDIR spectroscopy | Utilizes a quantum cascade laser to rapidly scan large sample areas and collect IR spectra and images. | **Particle Count:** Yes – automated imaging yields particle count, size, and shape.<br>**Mass Concentration:** Indirect – mass can be estimated from particle dimensions and density after calibration. | **High throughput & automation:** Enables rapid screening over large areas (110).<br>- Provides integrated morphological and chemical information. | **Detection limit:** Typically effective for particles >20 µm, possibly missing smaller particles (109,111).<br>- Narrower spectral range compared to conventional FTIR. |
| Py-GC/MS | Thermally decomposes samples into volatile pyrolysis products, which are then separated by GC and identified/quantified by MS. | **Particle Count:** No – quantifies the total mass of the polymer present but does not preserve physical particle information.<br>**Mass Concentration:** Yes – directly measures the mass concentration of polymers, regardless of particle size. | **Size-independent quantification:** Not limited by particle dimensions; ideal for micro- and nanoplastics.<br>- Provides accurate mass concentration data in complex matrices. | **Destructive:** The sample is completely degraded, losing count, size, and shape data.<br>- Susceptible to matrix interferences (e.g., lipids and other biomolecules may produce similar pyrolysis products as some plastics), requiring extensive quality control (80,112). |

Additionally, differences in quality control rigor, digestion methods, reference standards and spectral matching strategies further exacerbate differences in levels of MNPs detected. Although many studies emphasize plastic-free protocols during sample processing, other essential quality control measures such as whole-sample blank controls, spiking recovery experiments, and determination of detection and quantification limits are often inconsistently applied. In terms of digestion methods, use of acids, for example, has been shown to cause degradation of PA and discoloration of other types of MNPs (117).

#### D3: Bias in selection of participants into the study (or analysis)

The risk of selection bias was not significant, as researchers employed strict inclusion and exclusion criteria during subject recruitment. Many studies excluded participants with pre-existing comorbidities that could put them at risk of negative health outcomes, independent of their exposure to MNPs. However, many studies were also done in a single/dual-centre context. Four studies also recruited participants based on volunteerism, and ten out of 25 studies lacked control groups. The limited pool of participants and small sample sizes may impact the power of our studies and reduce the generalisability of the results (118).

#### D4: Bias due to post-exposure interventions

The risk due to post-exposure interventions in our studies is very low. Fourteen out of 25 studies were cross-sectional. Outcome measurement occurred at one time point and subjects were not followed up over time (119). The prospective study by Marfella et al. had a clearly defined end point, follow up of the participants stopped when a negative cardiovascular event was recorded. The preferential use of cross-sectional study design reflects the many challenges of performing prospective cohort studies in human subjects, including but not limited to ethical concerns of non-maleficence, difficulty in controlling exposure over time, and loss to follow up (119,120).

#### D5: Bias due to missing data

We noticed little evidence of missing data in the studies included. Most authors provided robust data on study population demographics, exposures and outcomes. This could be attributed to the retrospective and cross-sectional study design, where data could be easily retrieved, or only needed to be collected at one time point respectively. Many of the studies were also single/dual-centre, and cohort sizes were also relatively small. Nevertheless, bias arising from missing data will continue to be an important consideration in future research, when larger-scale, prospective studies are carried out.

#### D6: Bias arising from measurement of the outcome

We also assessed bias in this domain to be very low. Researchers ensured that methods of outcome measurements were the same for all participants, regardless of their exposure. Validated scoring systems to grade the severity of the health outcomes, such as the Nasal Obstruction Symptom Evaluation (NOSE) scale for rhinosinusitis in the study by Ta□ et al. and the APGAR score for newborn status in the study by Amereh et al. (22,24). However, more work needs to be done to standardise the biomarkers that are used as a reflection of specific pathological changes. Tailoring biomarkers to particular tissues and conditions enhances the precision of assessments and deepens our understanding of the role of MNPs in disease processes.

#### D7: Bias in selection of the reported result

Bias in this domain arises when study authors select results from a multiplicity of analyses based on the magnitude, and direction of *p-*value of the result. In our review, we noted that studies favoured more detailed reporting and discussion of health outcomes that showed a negative association with MNP exposures. With the exception of Nugrajapraha et al., reports also often glossed over non-significant correlations with specific disease markers, and the lack of a dose-response curve in many analyses. We also noted some preferential reporting of certain *p*-values, Rsquared coefficient and confidence intervals, based on whichever statistical method yielded more significant findings for the particular health outcome.

#### Overall judgement

We note that the studies included in this analysis were at highest risk of bias arising from uncontrolled confounding, lack of standardization and rigor in the measurement of MNP exposure, and selective reporting of outcome. These limitations temper confidence in observed associations and underscore the urgency for prospective, rigorously designed human studies with harmonized MNP quantification and comprehensive confounder adjustment.

## CONCLUSION

Our systematic review reveals that human *in vivo* evidence linking MNPs to adverse health outcomes is still nascent but growing rapidly. The most robust data emerge from cardiovascular and reproductive tissues, where PE, PP, PVC, PET, PS, and PA66 predominate. These MNPs appear to accumulate in vascular lesions, tumors, and reproductive fluids, with preliminary associations to inflammation, coagulation dysregulation, sperm dysfunction, and adverse obstetric outcomes. For the gastrointestinal and respiratory systems, emerging evidence suggests pro-inflammatory and microbiome-modulating effects. However, the overall quality of evidence is hindered by small sample sizes, cross-sectional designs, and methodological variability in MNP detection.

Before definitive public health recommendations can be made, larger, well-powered, longitudinal cohort studies with standardized exposure assessment protocols are needed. Specifically, future research should:

1. **Quantify MNP burdens prospectively** in blood and target tissues, incorporating standardized methods for MNP analysis and rigorous quality control to ensure reliability and reproducibility.
2. **Link MNP concentrations to clinical endpoints** in larger and more diverse cohorts.
3. **Adjust for confounding factors**—dietary habits, BMI, comorbidities, occupational exposures—to establish stronger causal inferences.
4. **Investigate mechanistic pathways** in humans, leveraging biomarkers of inflammation, oxidative stress, and immune activation.

## LIMITATIONS

The heterogeneity of study designs, MNP detection methods, and outcome measures precluded quantitative meta-analysis. Our analysis of the sources of MNP exposure was also limited by insufficient data in 13 out of 25 papers. Finally, selective reporting and incomplete adjustment for confounders in the included studies may overestimate the strength of observed associations.

## Supporting information

Supplementary Materials, including Full Search Terms

## Data Availability

Most data extracted and analyzed in this review, including study characteristics, risk-of-bias assessments, and search strategy, are available in the manuscript and supplementary materials. Additional de-identified summary data, including detailed polymer type information and the full data dictionary, will be made available upon reasonable request to the corresponding author.

## Contributors

AFWH conceived and designed the study. HAAT, DHT, and CTY performed the literature search. HAAT, DHT, CTY, and JJN performed the data extraction and risk-of-bias assessments, conducted the data synthesis, drafted the figures and tables, and wrote the first draft of the manuscript with input from all co-authors. All authors had full access to the underlying data; All authors reviewed and approved the final version and take responsibility for the decision to submit for publication.

## Declaration of interests

We declare no competing interests.

## Data sharing

Most data extracted and analyzed in this review—including study characteristics, risk-of-bias assessments, and search strategy—are available in the manuscript and supplementary materials. Additional de-identified summary data, including detailed polymer type information and the full data dictionary, will be made available upon reasonable request to the corresponding author. Access will be granted to researchers submitting a methodologically sound proposal and agreeing to a data-access agreement.

## Acknowledgments

We thank Ms Rebecca David for assistance with database search strategy development.

## Notes

### Competing Interest Statement

The authors have declared no competing interest.

### Clinical Protocols

https://www.crd.york.ac.uk/PROSPERO/view/CRD420251089766

