## Supplementary Materials, including Full Search Terms for "Health Impacts of Micro- and Nanoplastics in Humans: Systematic Review of *In Vivo* Evidence"

**Supplementary Table 1. Search strategies across databases.** **1**
**Supplementary Figure 1. Risk of Bias Assessment** **4**
**Supplementary Figure 2. Summary of risk-of-bias across studies by ROBINS-E domain.** **5**
**Supplementary Table 2. List of Articles Excluded at Full-Text Screening** **6**

**Supplementary Table 1. Search strategies across databases.**

| Database | Search Terms | Limits Applied | Results Retrieved |
|---|---|---|---|
| PubMed | (((“humans”[MeSH Terms] OR “infant”[MeSH Terms] OR “persons”[MeSH Terms] OR “adult”[MeSH Terms] OR “adolescent”[MeSH Terms] OR “aged”[MeSH Terms] OR “middle aged”[MeSH Terms] OR “child”[MeSH Terms] OR “young adult”[MeSH Terms])) AND (“microplastics”[MeSH Terms] OR “microplastics”[All Fields] OR (“plastic”[All Fields] AND “microparticles”[All Fields]) OR “plastic microparticles”[All Fields] OR “nanoplastics”[All Fields] OR “nanoplastic”[All Fields])) AND (“treatment outcome”[MeSH Terms] OR “health status”[MeSH Terms] OR “adverse effects”[MeSH Subheading] OR “toxicity”[MeSH Subheading] OR “risk assessment”[MeSH Terms] OR “public health”[MeSH Terms])) NOT (review[Publication Type] OR editorial[Publication Type] OR comment[Publication Type] OR letter[Publication Type]) | Humans, Excluded reviews, editorials, letters | 2199 |
| SCOPUS | TITLE-ABS-KEY ( "humans" OR "individuals" OR "persons" OR "adult" OR "adults" OR "elderly" OR "aged" OR "young adults" OR "children" OR "infant" OR "child" OR "adolescent" OR "middle-aged" ) AND TITLE-ABS-KEY ( "microplastics" OR "microplastic pollution" OR "plastic microparticles" OR "plastic nanoparticles" OR "nanoplastics" OR "plastic microbeads" ) AND TITLE-ABS-KEY ( "treatment outcome" OR "health status" OR "public health" OR "risk assessment" OR "adverse effects" OR "side effects" OR "toxicity" OR "health impact" OR "toxicology" OR "fatal outcome" OR "medical outcomes" OR "environmental risk" OR "health risk" ) | Subject area: Medicine; Excluded document types: reviews, editorials, letters, notes | 561 |

| Database | Search Terms | Limits Applied | Results Retrieved |
| --- | --- | --- | --- |
| Embase | 1. young adult/ or adult/ or human/ or infant/ or adolescent/ or aged/ or child/<br>2. microplastic/ or nanoplastic/ or plastic nanoparticles.mp.<br>3. treatment outcome/ or health status/ or functional status/ or health status indicator/ or fatality/ or clinical outcome/ or outcome assessment/ or patient-reported outcome/ or treatment failure/ or adverse outcome.mp. or risk assessment/ or health risk assessment/ or environment/ or public health/ or community medicine/ or preventative medicine/ or public health problem/ or toxic substance/ or toxicity/<br>4. 1 and 2 and 3<br>5. not (review or editorial or comment or letter or conference or interview).pt.<br>6. Final: 4 not 5 | Excluded reviews, editorials, comments, letters, conferences, interviews | 415 |
| Cochrane | #1: (humans OR infant OR persons OR adult OR adolescent OR aged OR "middle aged" OR child OR "young adult"):ti,ab,kw<br>#6: microplastic or microplastics or plastic microparticles or plastic nanoparticles or nanoplastic or nanoplastics or plastic microbeads<br>#7: treatment outcome or health status or health indicators or public health or health risk assessment or risk assessment or patient outcome or adverse effects or side effects or toxic effects or toxicity or toxicity assessment or long term effects or health impact or risk factors or adverse health outcomes or toxicology or environmental health or preventive medicine or risk management or fatal outcome or medical outcomes or health surveillance or toxicity or toxic effects or environmental risk or health risk assessment or toxic actions or public health assessment or health risks<br>#8: #1 AND #6 AND #7 | No filters applied; search limited to title, abstract, keywords | 22 |

| Database | Search Terms | Limits Applied | Results Retrieved |
|---|---|---|---|
| Web of Science | #1: TS=(young adult OR adult OR human OR infant OR adolescent OR aged OR child)<br>#2: TS=(microplastic OR nanoplastic OR plastic nanoparticles)<br>#3: TS=(microplastic OR nanoplastic OR plastic nanoparticles) AND Microplastics AND Nanoplastics AND Microplastic Particles<br>#4–5: Various combinations of TS=(treatment outcome OR health status OR health status indicator OR fatality OR clinical outcome OR outcome assessment OR patient-reported outcome OR treatment failure OR environment OR public health OR health risk assessment)<br>#6: #4 AND #2 AND #1<br>#7: #6 NOT DT=(Review) NOT DT=(Editorial Material) NOT DT=(Correction) NOT DT=(Letter) | Document Types: Excluded reviews, editorials, corrections, letters | 2347 |

Database-specific search strategies used in the systematic review, including search terms, applied limits, and number of records retrieved. Searches were conducted across PubMed, SCOPUS, Embase, Cochrane, and Web of Science, with filters applied to exclude non-original research articles such as reviews, editorials, letters, and commentaries where applicable.

**Supplementary Figure 1. Risk of Bias Assessment**

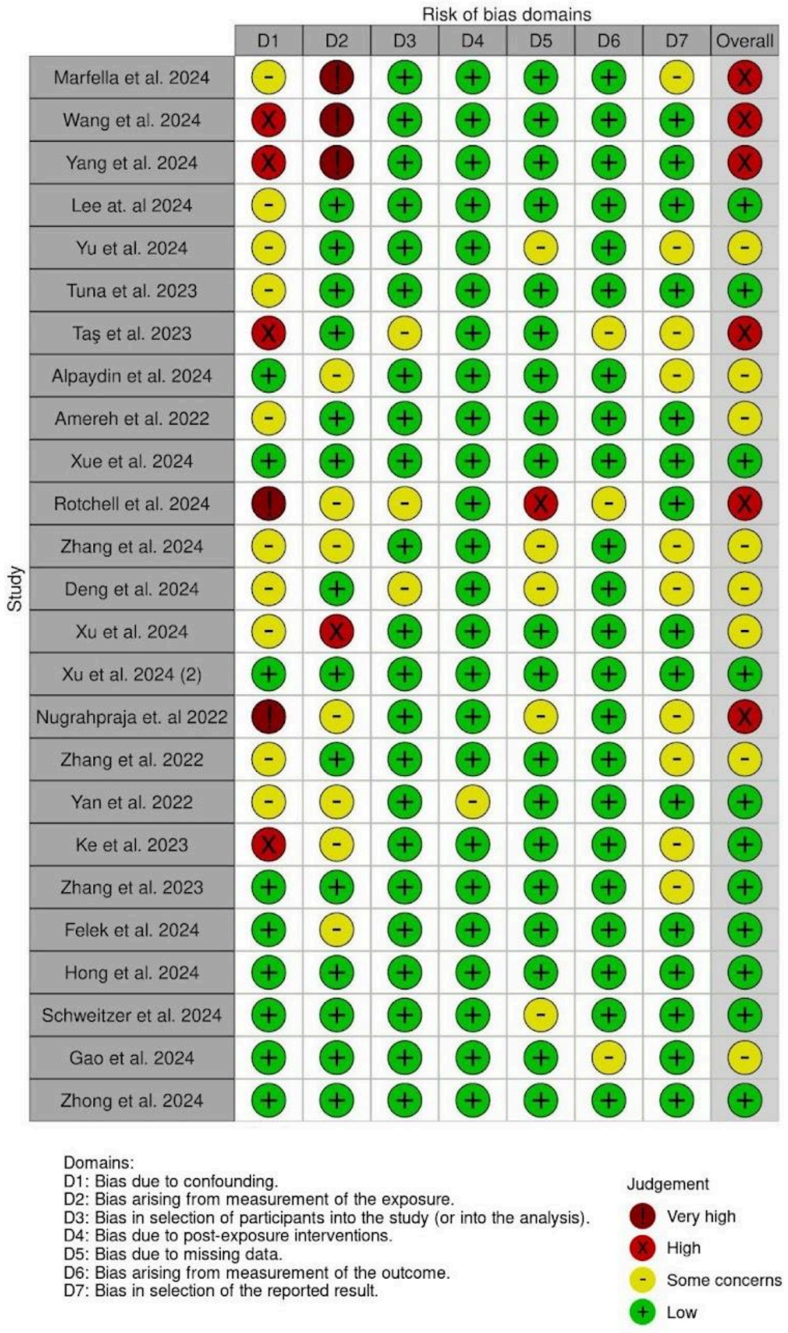


Heatmap summarising risk-of-bias judgments across seven ROBINS-E domains for each of the 25 human *in vivo* studies. Domains are: D1, confounding; D2, exposure measurement; D3, participant selection; D4, post-exposure interventions; D5, missing data; D6, outcome measurement; and D7, selective reporting. Symbols indicate domain-level risk: very high risk (dark red); high risk (red); some concerns (yellow); low risk (green). The "Overall" column shows each study's maximum domain rating, guiding interpretation of methodological robustness.

**Supplementary Figure 2. Summary of risk-of-bias across studies by ROBINS-E domain.**

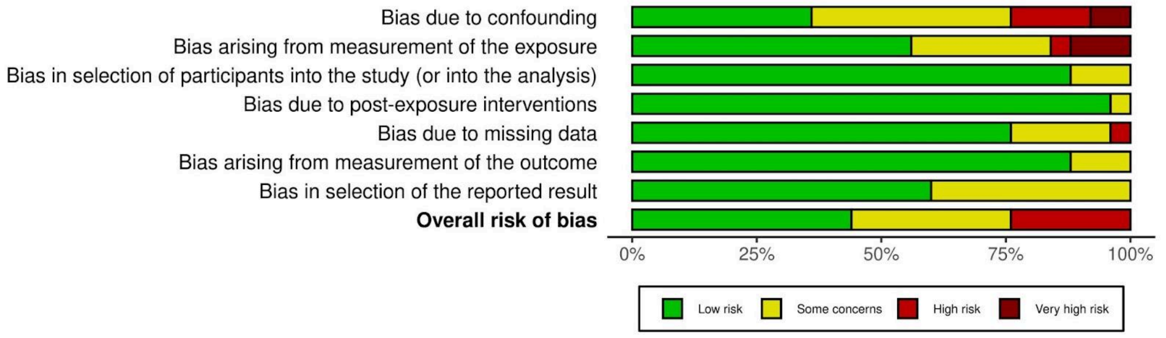


Stacked bar charts showing the proportion of included human *in vivo* studies rated as low risk (green), some concerns (yellow), high risk (red), and very high risk (dark red) within each ROBINS-E domain: D1 (confounding), D2 (exposure measurement), D3 (participant selection), D4 (post-exposure interventions), D5 (missing data), D6 (outcome measurement), D7 (selective reporting), and the overall risk of bias. Percentages along the x-axis reflect the share of studies falling into each risk category per domain

**Supplementary Table 2. List of Articles Excluded at Full-Text Screening**

| Articles excluded at Full-Text Screening | Author, Year of Publication | Reason for exclusion |
|---|---|---|
| Microplastics identified in plaques | Deutsche Apotheker Zeitung 2024 | Article is a news article, not original research |
| Immunotoxic, genotoxic, and endocrine-disrupting impacts of polyamide microplastic particles and chemicals | Alijagic et al. 2024 | Study used human cell models (US OS cell cultures). The microplastic particles were added ex-vivo. |
| An explorative study on respiratory health among operators working in polymer additive manufacturing. | Almstrand et al. 2023 | Research focused on ultrafine particles and volatile organic compounds, not microplastics specifically. |
| Are microplastics spreading infectious disease? | Beans 2023 | Article is a review / editorial, not original research |
| Effect of micro- and nanoplastics on the gastrointestinal mucosa and intestinal microbiome | Besednova et al. 2023 | Article is a review, not original research. It also focused on analysing animal studies and in-vitro models |
| Chronic exposure to polystyrene microplastics increased the chemosensitivity of normal human liver cells via ABC transporter inhibition. | Chen et al. 2024 | Study looked at effects of microplastics on THLE-2 cells in vitro. No in vivo experiments were carried out |
| Occurrence, toxicity and removal of polystyrene microplastics and nanoplastics in human sperm | Chen et al. 2024 | MP exposure was not natural; the sperm was incubated with polystyrene at 500 ug/ml in vitro which is 100 times more than detected normally in human semen |
| Environmental microplastics and nanoplastics: Effects on cardiovascular system | Chowdhury et al. 2024 | Article is a review, not original research |
| Polystyrene microplastics exposition on human placental explants induces time-dependent cytotoxicity, oxidative stress and metabolic alterations. | decSousa et al. 2024 | Study is done ex vivo, placenta was cultured with concentrations of microplastics outside of the body |
| Global trends and hotspots of gastrointestinal microbiome and toxicity based on bibliometrics | Duan et al. 2023 | Article is a systematic review, not original research. Furthermore, alongside microplastics, they also looked at the effects of drugs and other environmental pollutants on gut microbiome |
| Consumption of Bottled Water and Chronic Diseases: A Nationwide Cross-Sectional Study. | Dolcini et al. 2024 | Study has weak linkage and causation. Study also did not verify that the plastic bottles consumed contained microplastics |
| Examining the impact of nanoplastics and PFAS exposure on immune functions through inhibition of secretory immunoglobin A in human breast milk. | Enyoh et al. 2023 | Nanoplastics artificially introduced after the milk is taken out of the body |

| | | |
|---|---|---|
| Evaluation of nanoplastics toxicity to the human placenta in systems. | Enyoh et al. 2023 | Nanoplastics were artificially docked on to placental enzyme molecules in vivo. This study is a simulation model of human placenta. |
| Nanopolystyrene translocation and fetal deposition after acute lung exposure during late-stage pregnancy | Fournier et al. 2020 | Study looked at effects of nanopolystyrene pulmonary exposure in mice, not humans. |
| The male reproductive toxicity after nanoplastics and microplastics exposure: Sperm quality and changes of different cells in testis | Gao et al. 2023 | Study looked at the effects of sperm quality and testicular cells in mice, not humans |
| Microplastics and additives in patients with preterm birth: The first evidence of their presence in both human amniotic fluid and placenta | Halfar et al. 2023 | Study is mainly a detection study for presence of microplastics in amniotic fluid, does not have measurable health outcomes |
| Plastic in Arteries Tied With Higher Risk of Cardiovascular Problems. | Harris et al. 2024 | Short section in a news article, not original research |
| MRI-based microplastic tracking in vivo and targeted toxicity analysis | Hou et al. 2024 | Study looked at effects of microplastics in KM mice, not humans |
| Microplastics detected in cirrhotic liver tissue | Horvatits et al. 2024 | This is a “proof of concept” cause series and does not elaborate on any pathogenesis of microplastics. |
| Biological interactions of polystyrene nanoplastics: Their cytotoxic and immunotoxic effects on the hepatic and enteric systems | Huang et al. 2023 | Study involves cell lines in mice (AML-12) and human livers (L02). The section of the study done in vivo was done on mice. |
| Microplastic presence in dog and human testis and its potential association with sperm count and weights of testis and epididymis. | Hu et al. 2024 | The study only measured the clinical outcome relationships (IE testes weight and sperm count) in the dog testicles. |
| Toxicity of nanomixtures to human macrophages: Joint action of silver and polystyrene nanoparticles | Ilic et al. 2022 | Study involves human macrophage cell lines (THP-1). It also measures the effects of compounded silver and polystyrene nano particles, outside our scope of research |
| Vasomotor dysfunction in human subcutaneous arteries exposed ex vivo to food-grade titanium dioxide. | Jensen et al. 2018 | Human arteries were exposed to titanium dioxide ex-vivo. Titanium dioxide is also not a MNP. |
| Maternal nanoplastic ingestion induces an increase in offspring body weight through altered lipid species and microbiota | Jeong et al. 2024 | Study measured outcome in mice |
| Realistic Nanoplastics Induced Pulmonary Damage via the Crosstalk of Ferritinophagy and Mitochondrial Dysfunction. | Ji et al. 2024 | Study measured health outcome in mice for in vivo aspect. |

| | | |
|---|---|---|
| Microplastic toxicity and the gut microbiome | Karim et al. 2022 | Article is a book chapter, not original research |
| Polystyrene nanoplastics promote the blood-brain barrier dysfunction through autophagy pathway and excessive erythrophagocytosis | Kim et al. 2025 | Study primarily involves the use of brain endothelial cell culture |
| The Role of the Urban Exposome in the Increasing Global Rates of Pediatric Inflammatory Bowel Disease | Kuenzig et al. 2022 | The article is a review and not original research. It also discusses factors beyond the scope of our review, such as nighttime light, noise, green and blue space. |
| Micro(nano)plastics pollution and human health: How plastics can induce carcinogenesis to humans? | Kumar et al. 2022 | Article is a systematic review, not original research. |
| Microplastics and Cardiovascular Diseases: Importance of Coexisting Environmental Pollutants | Lee et al. 2024 | Article is a review/commentary article. It also focuses on various contaminants associated with MNP and not the MNPs alone |
| Mitigation of polystyrene microplastic-induced hepatotoxicity in human hepatobiliary organoids through bile extraction. | Li et al. 2024 | Study primarily measures outcomes in lab developed human hepatobiliary organoids |
| Microplastics aggravates rheumatoid arthritis by affecting the proliferation/migration/inflammation of fibroblast-like synovial cells by regulating mitochondrial homeostasis. | Lihua et al. 2023 | Study primarily involves cell culture (fibroblast-like synovialcytes) |
| Type-specific inflammatory responses of vascular cells activated by interaction with virgin and aged microplastics. | Lomonaco et al. 2024 | Study artificially exposes the vascular smooth muscle cells to the polystyrene and polyethylene microplastics in vitro |
| Mimicking human ingestion of microplastics: Oral bioaccessibility tests of bisphenol A and phthalate esters under fed and fasted states. | López-Vázquez et al. 2022 | Study used in-vitro models like the versantvoort model and UBM fasted state model to measure health outcomes. |
| Relationship between microplastics and cardiovascular risk factors | Mamedov et al. 2024 | Article is a review, not original research. |
| Microplastics in human food chains: Food becoming a threat to health safety | Mamun et al. 2023 | Article is a review, not original research |
| Cellular response of keratinocytes to the entry and accumulation of nanoplastic particles | Martin et al. 2024 | Study involves. the use of various lab-developed models to simulate skin. |
| Implications of polystyrene and polyamide microplastics in the adsorption of sulfonamide antibiotics and their metabolites | Mejías et al. 2024 | Focus of the study is wrong. Study's primary aim is to look at the adsorption of sulfonamides, not MNPs looks at Study involves use of |

| | | |
|---|---|---|
| in water matrices | | |
| Microplastics and nanoplastics-A new cardiovascular risk factor | Münzel et al. 2024 | Article is a commentary on an included study by Marfella et al. |
| Toxicity of microplastic fibers containing azobenzene disperse dyes to human lung epithelial cells cultured at an air-liquid interface | O'Connor et al. 2024 | Study is in-vitro, uses human lung epithelial cells |
| Effect of altered human exposome on the skin and mucosal epithelial barrier integrity. | Pat et al. 2023 | Article is a review, not original research |
| Plastic exposure may be associated with the deposition of microplastics in reproductive tissues and adverse clinical outcomes | Phillipi et al. 2023 | Article is a systematic review, not original research |
| Environmental pollutant exposure can exacerbate COVID-19 neurologic symptoms | Reyes et al. 2020 | Study focuses primarily on particulate matters and environmental pollutants, instead of microplastics |
| Breathing in danger: Mapping microplastic migration in the human respiratory system | Riaz et al. 2024 | Study used numerical models, not humans |
| Contribution of Cancer-Specific Protein Coronas to the Pro-Tumor Effects of Nanoplastics through Enhanced Cellular Interactions. | Tang et al. 2024 | Study added NPs ex-vivo |
| Effects of polystyrene microplastics on the metabolic level of Pseudomonas aeruginosa. | Tao et al. 2024 | Study discusses the effect of microplastics on bacteria not humans |
| Microplastics accumulated in breast cancer patients lead to mitophagy via ANXA2-mediated endocytosis and IL-17 signaling pathway. | Tian et al. 2025 | Microplastics are added in vitro, after the removal of the breast cancer tissue. |
| Micro/Nanoplastic Exposure on Placental Health and Adverse Pregnancy Risks: Novel Assessment System Based upon Targeted Risk Assessment Environmental Chemicals Strategy | Wan et al. 2024 | Study does not give solid evidence to show any adverse health outcomes in patients. This paper is simply suggesting a future study design. |
| Implication of ferroptosis in hepatic toxicity upon single or combined exposure to polystyrene microplastics and cadmium | Wang et al. 2023 | Study involves cell lines and also measures outcomes on mice |
| Microplastics, cardiometabolic risk, genetics and Alzheimer's disease. | Watts et al. 2022 | Article is an editorial, not original research |
| Nanoplastics Penetrate Human Bronchial Smooth Muscle and Small Airway Epithelial Cells and Affect Mitochondrial Metabolism. | Winiaskar 2024 | Study is in vitro, and makes use of Bronchial Smooth Muscle and Small Airway Epithelial cell cultures |

| | | |
|---|---|---|
| Pigment microparticles and microplastics found in human thrombi based on Raman spectral evidence | Wu et al. 2023 | Particles found in study were not exactly focused on microplastics |
| Phthalates released from microplastics inhibit microbial metabolic activity and induce different effects on intestinal luminal and mucosal microbiota. | Yan et al. 2022 | Study used simulated intestinal microbiota models (M-SHIME) to stimulate the intestines |
| Understanding the impact of nanoplastics on reproductive health: Exposure pathways, mechanisms, and implications | Ye et al. 2024 | Article is a review, not original research |
| Multi-omics analysis reveals size-dependent toxicity and vascular endothelial cell injury induced by microplastic exposure in vivo and in vitro | Zhang et al. 2022 | Study involves rats in in vivo aspect and the microplastics exposure was not natural |
| Polystyrene microplastics disturb maternal glucose homeostasis and induce adverse pregnancy outcomes | Zhang et al. 2024 | Study measures outcomes in mice, not humans |

Summary of 56 article titles, authors, year of publication and reasons for exclusion at full-text screening stage.
